# Declining Course of Humoral Immune Response in Initially Responding Kidney Transplant Recipients after Repeated SARS-CoV-2 Vaccination

**DOI:** 10.1101/2022.03.29.22272858

**Authors:** Simon Ronicke, Bilgin Osmanodja, Klemens Budde, Annika Jens, Charlotte Hammett, Nadine Koch, Bianca Zukunft, Friederike Bachmann, Mira Choi, Ulrike Weber, Bettina Eberspächer, Jörg Hofmann, Fritz Grunow, Michael Mikhailov, Fabian Halleck, Eva Schrezenmeier

## Abstract

Immunogenicity of SARS-CoV-2 vaccines in kidney transplant recipients is limited, resulting in inadequately low serological response rates and low immunoglobulin (Ig) levels, correlating with reduced protection against death and hospitalization from COVID-19. We retrospectively examined the time course of anti-SARS-CoV-2 Ig antibody levels after up to five repeated vaccinations in 644 previously nonresponding kidney transplant recipients. Using anti SARS-CoV-2 IgG/IgA ELISA and the total Ig ECLIA assays, we compare antibody levels at 1 month with levels at 2 and 4 months, respectively. Additionally, we correlate the measurements of the used assays.

Between 1 and 2 months, and between 1 and 4 months, mean anti-SARS-CoV-2 Ig levels in responders decreased by 14% and 25%, respectively, depending on the assay. Absolute Ig values and time course of antibody levels and showed high interindividual variability. Ig levels decreased by at least 20% in 77 of 148 paired samples with loss of sufficient serological protection over time occurring in 18 out of 148 (12.2%).

IgG ELISA and total Ig ECLIA assays showed a strong positive correlation (Kendall’s tau=0.78), yet the two assays determined divergent results in 99 of 751 (13.2%) measurements. IgG and IgA assays showed overall strong correlation but divergent results in 270 of 1.173 (23.0%) cases and only weak correlation of antibody levels in positive samples.

Large interindividual variability and significant loss of serological protection after 4 months supports repeated serological sampling and consideration of shorter vaccination intervals in kidney transplant recipients.

## 1. Introduction

Vaccination against severe acute respiratory syndrome coronavirus 2 (SARS-CoV-2) induces a rapid and strong immunological response in healthy individuals (1). Anti-SARS-CoV-2 antibodies are a serological marker of an adequate immune response and correlate with protection against coronavirus disease 2019 (COVID-19) induced by vaccination (2). In particular, IgG antibodies correlate with protection from death and hospitalization due to COVID-19 (3,4). Two doses of vaccine usually induce sufficient antibodies for protection against the SARS-CoV-2 Alpha and Delta variant, whereas three doses are required to induce protection against the Omicron variant in healthy individuals (5).

Kidney transplant recipients (KTR) show a secondary immunodeficiency caused by the intake of immunosuppressive medication (6) and chronic kidney disease (7). Specifically, reduced immunogenicity of SARS-CoV-2 vaccines leads to a low rate of sufficient serological response – below 50% after the third dose of vaccine – and lower levels of antibodies in KTR (8–10). The result is a lack of protection against COVID-19 in KTR as compared with healthy individuals (11,12). A third vaccination was recommended early on for KTR in order to increase immune response (13). Further, repeated vaccinations under modulated immunosuppression effectively increase protection, yet a substantial number of patients does not reach protective antibody levels (14,15).

Vaccine effectiveness after two and three doses of vaccine vanishes over time even in healthy individuals, limiting the duration of protection (1). Whether the limited immune response in KTR leads to a faster reduction in protection is not known (15).

In the current study, we assess the course of anti-SARS-CoV-2 antibodies over time in KTR who show serological response after receiving two to five doses of SARS-CoV-2 vaccines. We evaluate the serological response with two different Ig assays. Finally, we correlate measurements between IgG and IgA assays.

## 2. Materials and Methods

Kidney transplant recipients treated and followed at our institution received repeated doses of SARS-CoV-2 vaccines in case of sustained non-response to vaccination against SARS-CoV-2 (14). Data from up to five doses of vaccine were included in this analysis. Basic immunization was performed with two doses; third, fourth and fifth immunizations were performed with one dose of BNT162b2 (Comirnaty, BioNTech/Pfizer), mRNA-1273 (Spikevax, Moderna Biotech), ChAdOx1-S (AZD1222, AstraZeneca) or Ad26.COV2.S (Johnson & Johnson, Janssen) in different combinations. We obtained written and informed consent into off-label use for vaccine doses four and five from all patients.

At routine visits, serological response following vaccinations was measured using different assays either alone or in parallel:

1. An anti-SARS-CoV-2 enzyme-linked immunosorbent assays (ELISA) for the detection of IgG antibodies against the S1 domain of the SARS-CoV-2 spike (S) protein in serum according to the instructions of the manufacturer (Anti-SARS-CoV-2-ELISA (IgG), EUROIMMUN Medizinische Labordiagnostika AG, Lübeck, Germany) (16). Processing and measurement were done using the fully automated "Immunomat” (Institut Virion\Serion GmbH, Würzburg, Germany). Results were determined by comparing the obtained signals of the patient samples with the previously obtained cut-off value of the calibrator. As suggested by the manufacturer, we considered samples with a cut-off index ≥1.1 positive for IgG and IgA.
2. An electrochemiluminescence immunoassay (ECLIA, Elecsys, Anti-SARS-CoV-2, Roche Diagnostics GmbH, Mannheim, Germany) for the detection of human immunoglobulins, including IgG, IgA and IgM against the spike receptor binding (RBD) domain protein. Results were determined by comparing the obtained signals of the patient samples with the previously obtained cut-off value of the calibrator. As suggested by the manufacturer and as recommended by Caillard et al (15), we considered samples with a cut-off index ≥ 264 U/ml positive.

The standard maximum level determined was >2500 U/ml. With regard to the following analyses, we defined the maximum measurement of >2500 U/ml as equal to 2500 U/ml and removed measurements that were only performed to a maximum dilution of >250 U/ml from the dataset.

We retrospectively analyzed serological response to all basic immunizations, third, fourth and fifth immunizations performed between December 27^th^ 2020 and December 31^st^ 2021. We included serological data of COVID-naïve and previously non-responding adult kidney transplant recipients who received at least one SARS-CoV-2 vaccination after kidney transplantation into the analysis. Conversely, any positive SARS-CoV-2 RNA PCR, positive anti-SARS-CoV-2-N-protein antibodies, positive anti-SARS-CoV-2 Ig or administration of monoclonal anti-SARS-CoV-2-S-protein antibody therapy before the serological sample lead to exclusion of the respective following serological data. Samples performed within less than 14 days after vaccination were not included.

The primary outcome was the course of serological response within the vaccination interval, hence after the respective vaccination and before any further vaccination. The secondary outcomes were the correlations of serological measurements between the two aforementioned anti-SARS-CoV-2 Ig ELISA (IgG) and ECLIA (total Ig) assays and the anti-SARS-CoV-2 IgA ELISA assay.

For the analysis of the course of anti-SARS-CoV-2 Ig after vaccination, we only included patients who showed a positive serological response to the respective vaccination. We assigned all serological samples to periods with regard to their time distance to the date of vaccination: First period at 2 to 6 weeks (14 to 41 days), second period at 6 to 12 weeks (42 to 83 days), and third period at 12 to 40 weeks (84 to 279 days) after vaccination. We evaluated the results of the two different anti-SARS-CoV-2 Ig assays separately. In case of multiple samples in the same patient within the same period in the same vaccination interval, we kept only the first sample. We paired the data from the first period with the data from the second and third period, respectively, in all patients with available anti-SARS-CoV-2 Ig samples in the respective pair of periods. Finally, we compared the level of anti-SARS-CoV-2 Ig at the first period with the level at the second and third period, respectively. We performed two-sided Wilcoxon signed rank tests to test for differences between the periods.

To correlate serological measurements between the two anti-SARS-CoV-2 Ig assays (IgG ELISA and IgG ECLIA) as well as between these Ig and the anti-SARS-CoV-2 IgA assay (IgA ELISA), we compared pairs of data that came from the same patient at the same date in graphical analysis. To determine statistical relationship between the assay’s results, we calculated Pearson product-moment correlation coefficient for parametric or Kendall rank correlation test for non-parametric data after testing for normality using Shapiro-Wilk normality test.

Statistical analysis R studio v. 1.4.1717 and R version 4.1.1 (2021-08-10) was used to perform the statistical analysis. We applied a significance level alpha = 0.05 for all calculations.

The institutional ethics committee of Charité – Universitätsmedizin Berlin approved this retrospective analysis (ethics votum EA1/030/22).

## 3. Results

8.409 serological samples after 2.799 SARS-CoV-2 vaccinations in 1.369 patients were initially evaluated. **Figure 1** illustrates the process of data exclusion and the split into datasets comprised of serological samples performed with the respective IgG ELISA, Ig ECLIA, and IgA ELISA assays. **Figure 1** also presents the amount of paired data used for the following comparison of anti-SARS-CoV-2 IgG levels in different periods and the selection of samples used for correlations between the assays. **Table 1** presents the baseline demographic and immunosuppression data of the 644 included patients and characteristics of the 925 included vaccinations.

**Figure 1.**
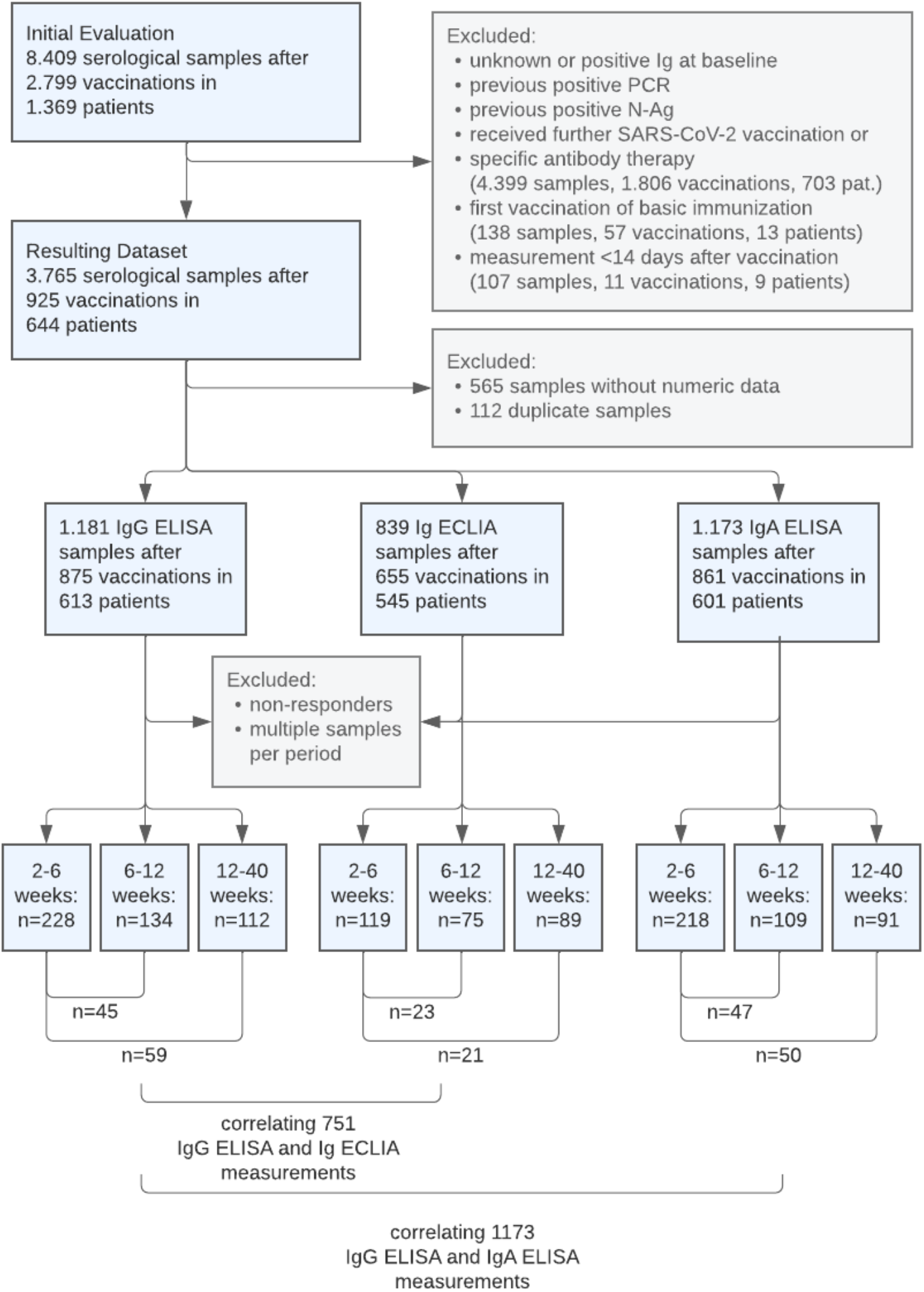
Flow diagram of evaluated, excluded, and included serological sample data.

**Table 1.**
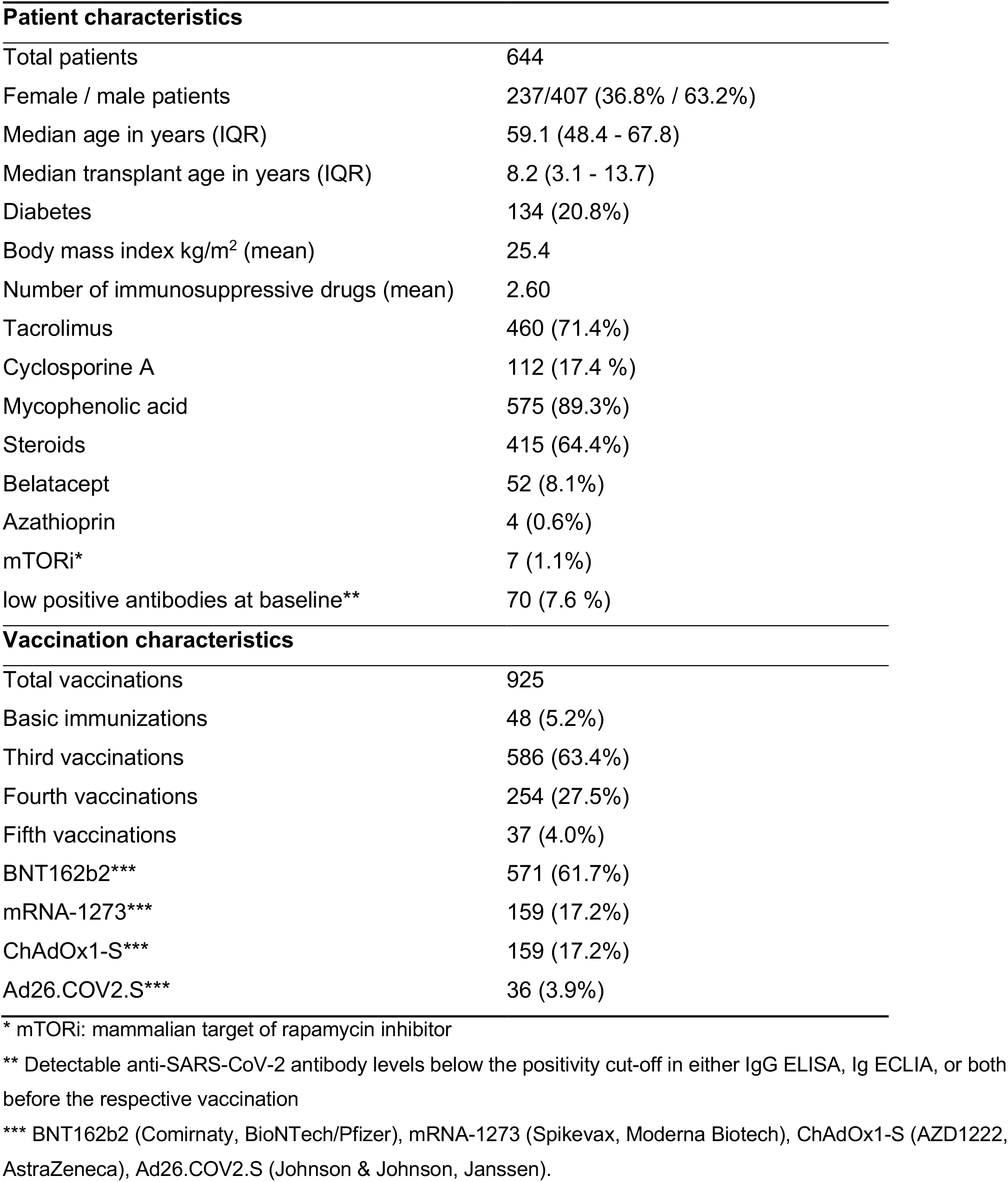
Baseline characteristics of the included patients and SARS-CoV-2 vaccinations.

Serological samples from the first, second, and third period were performed at a median of 32 days (1 month), 61 days (2 months) and 124 days (4 months), respectively, after the date of vaccination. In vaccine responders, mean Ig levels determined with ELISA and ECLIA showed a large variability. Mean IgG ELISA, Ig ECLIA and IgA ELISA levels peaked at 1 month and decreased by 14%, 25%, and 17%, respectively, essentially already at 2 months without substantial further decrease at 4 months (**Table 2**).

**Table 2.**
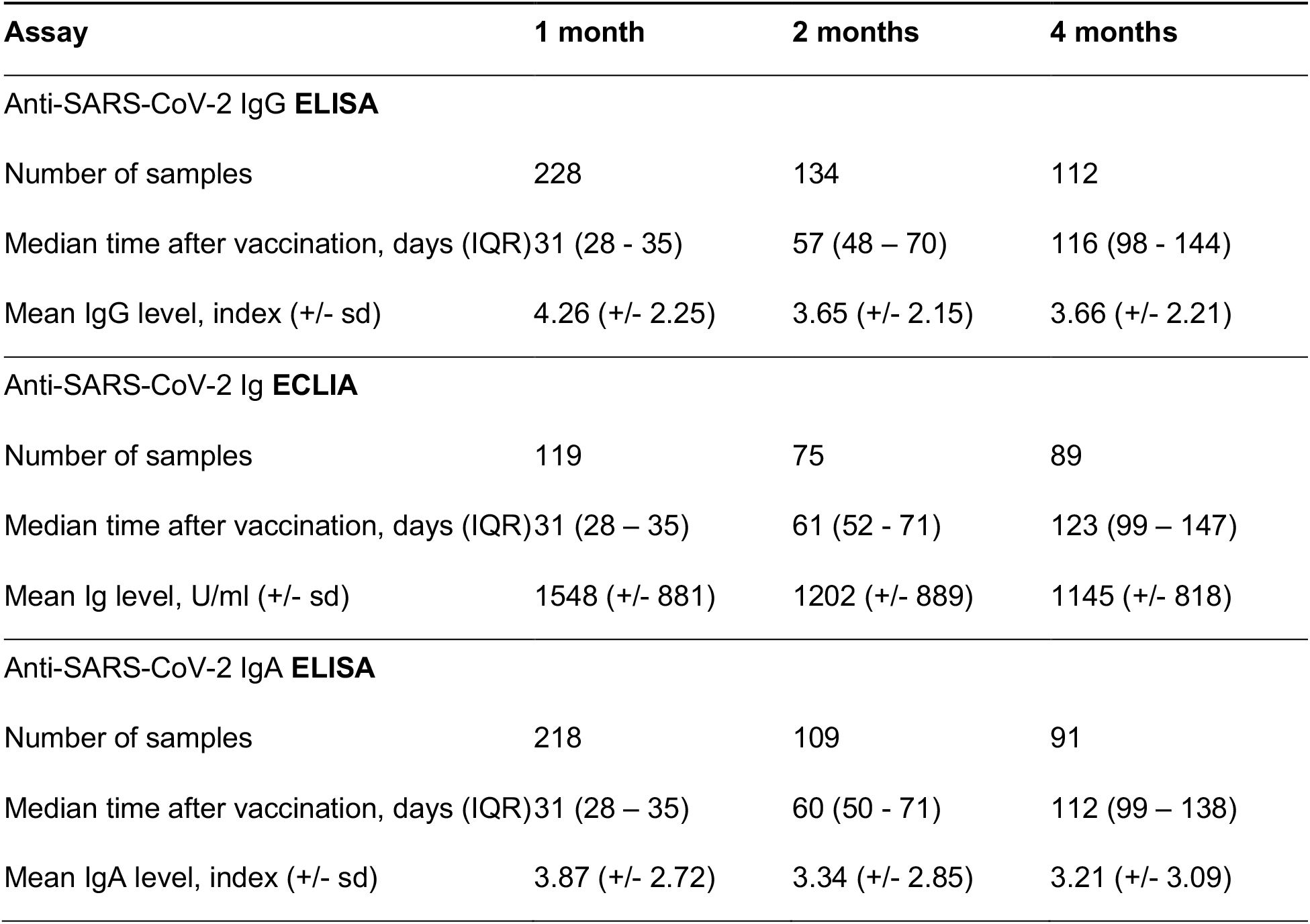
Mean anti-SARS-CoV-2 IgG, Ig and IgA levels in responders with samples at different intervals, relating to the respective IgG, Ig and IgA assays.

| **Assay** | **1 month** | **2 months** | **4 months** |
| --- | --- | --- | --- |
| **Anti-SARS-CoV-2 IgG ELISA** |  |  |  |
| Number of samples | 228 | 134 | 112 |
| Median time after vaccination, days (IQR) | 31 (28 - 35) | 57 (48 - 70) | 116 (98 - 144) |
| Mean IgG level, index (+/- sd) | 4.26 (+/- 2.25) | 3.65 (+/- 2.15) | 3.66 (+/- 2.21) |
| **Anti-SARS-CoV-2 Ig ECLIA** |  |  |  |
| Number of samples | 119 | 75 | 89 |
| Median time after vaccination, days (IQR) | 31 (28 - 35) | 61 (52 - 71) | 123 (99 - 147) |
| Mean Ig level, U/ml (+/- sd) | 1548 (+/- 881) | 1202 (+/- 889) | 1145 (+/- 818) |
| **Anti-SARS-CoV-2 IgA ELISA** |  |  |  |
| Number of samples | 218 | 109 | 91 |
| Median time after vaccination, days (IQR) | 31 (28 - 35) | 60 (50 - 71) | 112 (99 - 138) |
| Mean IgA level, index (+/- sd) | 3.87 (+/- 2.72) | 3.34 (+/- 2.85) | 3.21 (+/- 3.09) |

### 3.1 Course of anti-SARS-CoV-2 Ig after SARS-CoV-2 Vaccination in paired samples

The specific comparison of anti-SARS-CoV-2 Ig levels of responders at 1 month with levels at 2 months and 4 months in paired samples from both IgG ELISA and Ig ECLIA assays showed decreasing Ig levels in 77 of 148 (52.0%) of vaccination cases, increasing Ig levels in 26 of 148 (17.6%) cases and stable Ig levels (+/-20%) in the remaining 45 cases (30.0%) (**Figure 2**). In 18 out of 137 paired samples (12.2%) with positive Ig levels at 1 month, Ig levels were determined below the respective cut-off at a later period.

**Figure 2.**
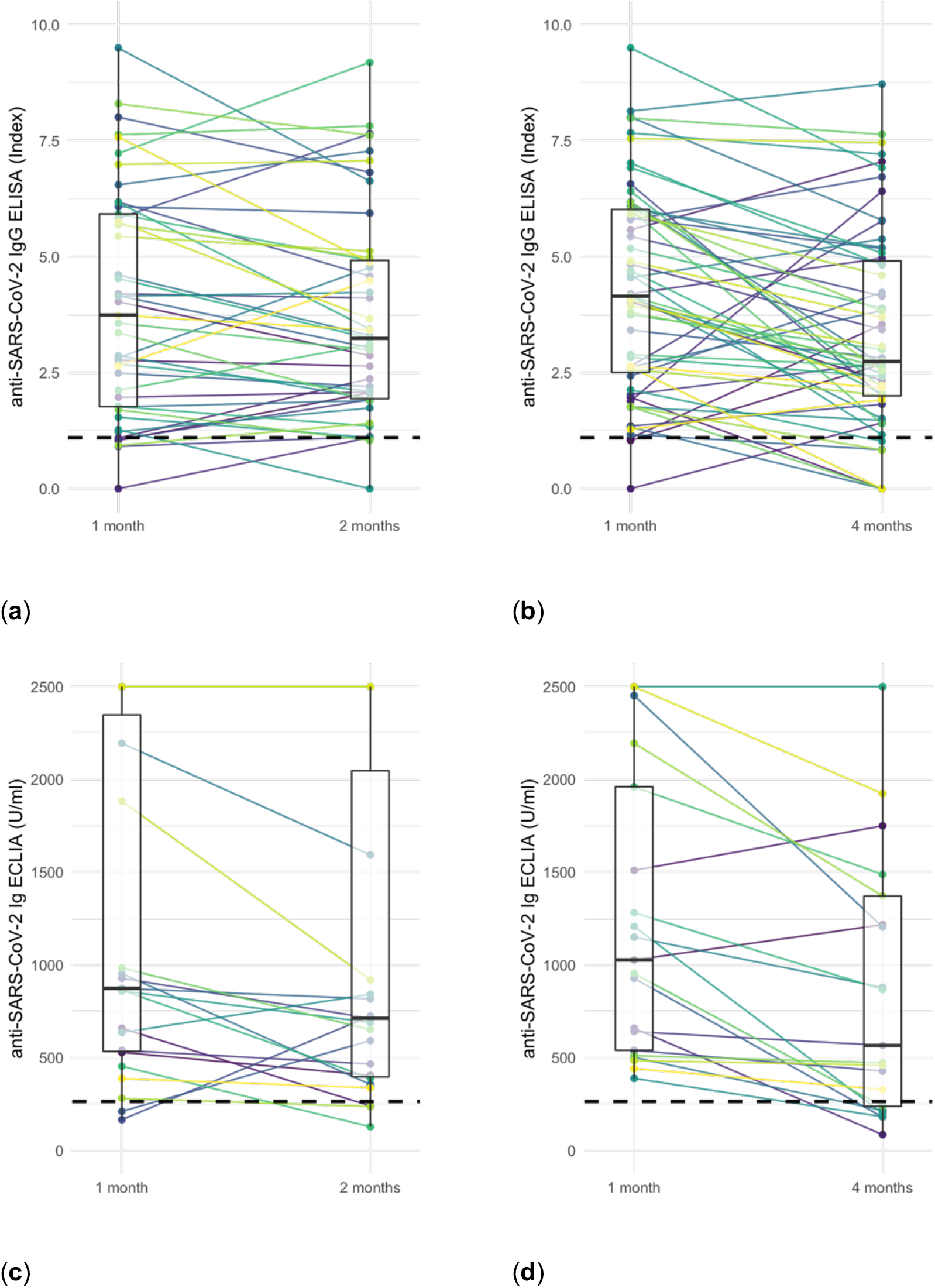
Comparison of anti-SARS-CoV-2 Ig levels at different intervals based on ELISA IgG and ECLIA Ig assays: **(a)** ELISA at 1 month vs. 2 months, **(b)** ELISA at 1 month vs. 4 months, **(c)** ECLIA at 1 month vs. 2 months, and **(d)** ECLIA at 1 month vs. 4 months. Wilcoxon signed rank test showed a significant decrease between 1 and 4 months (ELISA: p < 0.001, ECLIA: p = 0.005). Dashed line: positivity cut-off at 1.1 index or 264 U/ml, respectively. Boxplots’ line, lower and upper hinge: median, first and third quartile.

The decrease of anti-SARS-CoV-2 Ig levels between 1 and 4 months in both ELISA and ECLIA Ig assays was significant according to two-sided Wilcoxon signed rank test (ELISA: p < 0.001, ECLIA: p = 0.005). The comparison of anti-SARS-CoV-2 Ig levels between 1 and 2 months, however, was only significant in ECLIA samples but not in ELISA (ELISA: p = 0.12, ECLIA: p < 0.05).

Responders with increasing Ig levels after vaccination were more frequently observed to be younger, non-diabetic and receiving mycophenolic acid and belatacept immunosuppression (**Table 3**).

**Table 3.** Comparison of baseline characteristics of patients with anti-SARS-CoV-2 Ig levels increasing vs. decreasing at least 20% from 1 to 2 or 4 months after vaccination in paired samples.

| Patient characteristics | Increasing Ig level | Decreasing Ig level |
| --- | --- | --- |
| Number of patients | 22 | 47 |
| Female / male patients | 15/7 (68% / 32%) | 26/21 (55% / 45%) |
| Median age in years (IQR) | 41.1 (33.3 – 54.4) | 60.2 (49.5 – 71.1) |
| Median transplant age in years (IQR) | 5.7 (3.7 – 8.5) | 7.0 (3.4 – 11.2) |
| Diabetes | 3 (13.6%) | 11 (23.4%) |
| Body mass index (mean) | 24.5 | 25.5 |
| No. of immunosuppressives (mean) | 2.5 | 2.2 |
| Tacrolimus | 17 (77.3%) | 35 (74.5%) |
| Cyclosporine A | 2 (9.1 %) | 9 (19.1 %) |
| Mycophenolic acid | 21 (95.5%) | 21 (44.6%) |
| Steroids | 11 (50.0%) | 37 (78.7%) |
| Belatacept | 2 (9.1 %) | 2 (4.2%) |

### 3.2. Correlation of anti-SARS-CoV-2 IgG ELISA and Ig ECLIA

Correlation of 751 anti-SARS-CoV-2 IgG ELISA with Ig ECLIA assay results showed a strong positive association between the two tests (**Figure 3**) (Kendall’s tau-b correlation coefficient = 0.784, p < 0.001). Despite the strong positive correlation, we observed cases that were positive in ELISA and negative in ECLIA (97 of 751; 12.8%) and vice versa (2 of 751; 0.2%) (**Table 4**).

**Figure 3.**
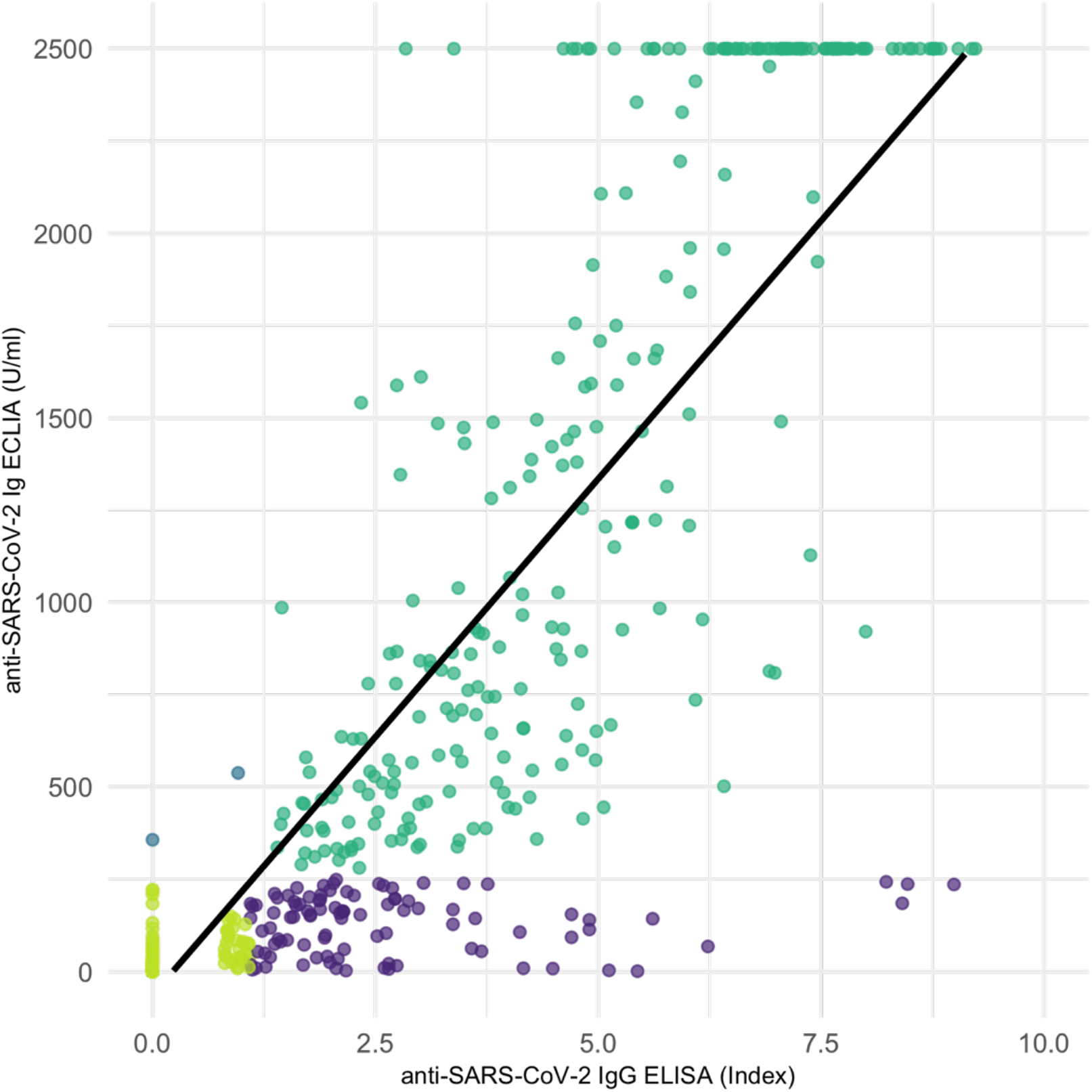
Correlation scatter plot of 751 serological samples performed with anti-SARS-CoV-2 IgG ELISA and Ig ECLIA assays showing a strong positive association in Kendall rank correlation test (tau = 0.784, p < 0.001). Violet: 97 samples determined positive in ELISA but negative in ECLIA. Blue: Two samples determined positive in ECLIA but negative in ELISA. Dark green and light green: congruent results determined as positive or negative, respectively. Black line: linear correlation.

**Table 4.** Correlation table of 751 anti-SARS-CoV-2 IgG ELISA and Ig ECLIA assay results.

| Assay | IgG ELISA |  |  |  |
| --- | --- | --- | --- | --- |
|  |  | positive | negative | total |
| Ig ECLIA | positive | 260 | 2 | 262 |
|  | negative | 97 | 392 | 489 |
|  | total | 357 | 394 | 751 |

### 3.3. Course of anti-SARS-CoV-2 IgA after SARS-CoV-2 Vaccination in paired samples

Anti-SARS-CoV-2 IgA levels of IgA-responders at 1 month with levels at 2 months and 4 months in paired samples from IgA ELISA assays showed decreasing IgA levels in 50 of 97 (51.5%) of cases, increasing IgA levels in 9 of 97 (9.2%) cases and stable Ig levels (+/-20%) in the remaining 38 cases (39.2%) (**Figure 4**). 21 of 97 (21.6%) became negative. The decrease of anti-SARS-CoV-2 Ig levels between 1 and 2 months as well as between 1 and 4 months was significant according to two-sided Wilcoxon signed rank test (p < 0.001 for both).

**Figure 4.**
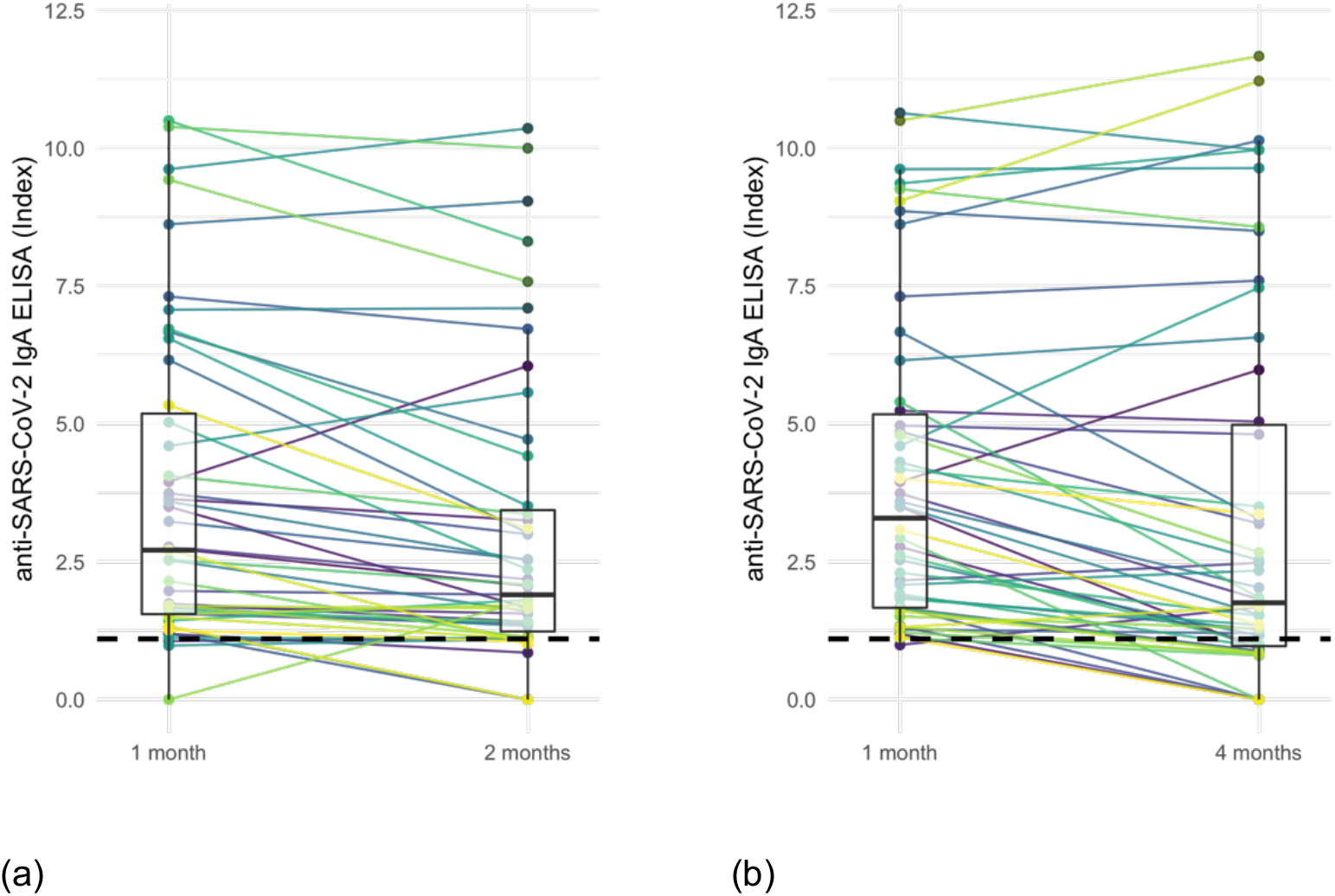
Comparison of anti-SARS-CoV-2 IgA levels at different intervals based on ELISA IgA assay: **(a)** IgA at 1 month vs. 2 months, **(b)** IgA at 1 month vs. 4 months. Wilcoxon signed rank test showed a significant decrease between 1 and 2 months (p < 0.001) as well as between 1 and 4 months (p < 0.001). Dashed line: positivity cut-off at 1.1 index. Boxplots’ line, lower and upper hinge: median, first and third quartile.

### 3.4. Correlation of anti-SARS-CoV-2 IgG and IgA

Correlation of all 1.173 anti-SARS-CoV-2 IgG ELISA with corresponding IgA ELISA assay results showed a moderate positive association between the two variables (**Figure 5**) (Kendall’s tau-b correlation coefficient = 0.499, p < 0.001). However, IgA and IgG assays determined differing results in 266 of 1.173 cases (**Table 4**). In cases with divergent results, the percentage of IgG only vs. IgA only positive measurements increased with time after the vaccination (supplement figure 1 and 2).

**Figure 5.**
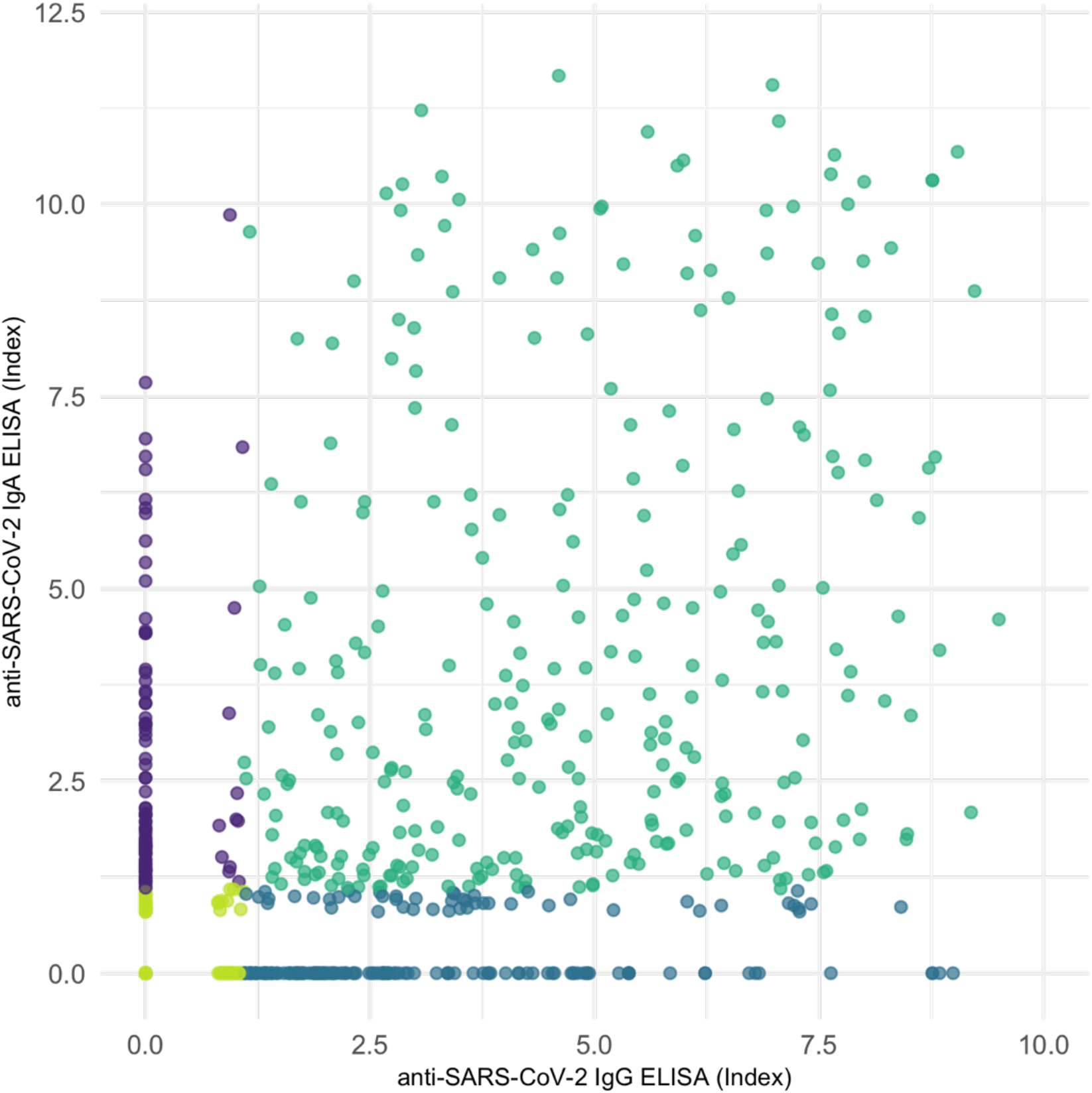
Correlation scatter plot of serological samples performed with anti-SARS-CoV-2 IgG ELISA and IgA ELISA assays. Correlation shows a moderate positive association in Kendall rank correlation test (tau = 0.499, p < 0.001). Violet: samples determined positive in IgA but negative in IgG assays. Blue: samples determined negative in IgA but positive in IgG assays. Dark green and light green: congruent results determined as positive or negative, respectively.

We found only a weak correlation when limiting the analysis to data pairs with positive measurements in at least one (tau = 0.141, p < 0.001) or in both of the compared IgG and the IgA assay (tau = 0.176, p < 0.001).

In both assays, variable distribution did not follow a normal distribution according to Shapiro-Wilk normality test.

**Table 4.** Correlation table of anti-SARS-CoV-2 IgG ELISA and IgA ELISA samples.

| Assay | IgG ELISA |  |  |  |
| --- | --- | --- | --- | --- |
|  |  | positive | negative | total |
| IgA ELISA | positive | 298 | 96 | 394 |
|  | negative | 174 | 605 | 779 |
|  | total | 472 | 701 | 1.173 |

## 4. Discussion

The current study presents the first systematic analysis of anti-SARS-CoV-2 Ig antibody course after vaccination in a large cohort of previously nonresponding KTR who were subsequently receiving up to five doses of vaccine.

While it is known that rates of serological non-responders are inadequately high among KTR (9,13), we were able to show that levels of humoral protection decrease early also in responding KTR irrespective of the type of Ig and the assay used. Loss of antibodies at 2 months and 4 months after vaccination occurred in a substantial number of initially responding individuals, presumably resulting in a loss of protection against COVID-19. While it is known that patient receiving MPA have a low vaccination response rate (14,17), our results suggest that use of MPA is also associated with a delay in vaccination response resulting in increasing Ig levels over time in almost 18% of patients. Most importantly, we noted a high variability in the development of humoral protection over time between KTR. These results support the monitoring of antibody levels and, if required, shorter vaccination intervals in KTR compared with healthy individuals.

The loss of protective Ig levels over time is more pronounced in KTR than in healthy individuals who are able to generate a detectable immune response over a period of more than six months. (18,19) Contrary to patients receiving hemodialysis who show high response rates after two doses of vaccine, (20) KTR do require repeated doses to elicit a sufficient humoral response. Still, the decrease of mean Ig levels in both hemodialysis patients and KTR might be comparable. (20) Similar to our results, Weigert et al. report a 25% decrease in IgG from 42 to 140 days after vaccination. In both populations loss of protection does occur faster and more frequently than in healthy individuals.

It should be noted that the used Ig level cut-offs are indicative of protection against SARS-CoV-2 variants before the Delta variant. Although we cannot determine any clear cut-off values, loss of protection against Delta and Omicron variants will be more severe as these variants have been shown to require a higher level of humoral response for protection. (5) Even though humoral response correlates with protection from disease, specific neutralizing antibodies and T cell response are important factors that we did not cover in this analysis.

The correlation of two different anti-SARS-CoV-2 Ig assays, namely one IgG and one total Ig assay, in our population revealed that despite a strong positive correlation between the assays’ results, a relevant number of individuals presented with diverging results. This observation reflects some differences in sensitivity between both assays. Whether this should impact clinical practice and trigger the use of multiple assays, preference of one assay over the other, cannot be concluded from our analysis.

Correlation of IgG and IgA assay results indicates that these lead consistent interpretations of results in a majority of cases. However, in about a quarter of cases we observed diverging results, and moreover, the level of IgG and IgA antibodies did not correlate well in positive cases. In previous studies, KTR with positivity in either one of the assays were classified as responders, what might have led to an overinterpretation of vaccine response in KTR (10).

Due to the study’s retrospective design, methodological limitations arise. However, the applied selection criteria for sample data account for the risk of selection bias. In a large number of patients at our institution, repeated serological measurements were not performed thus resulting in exclusion of singular samples. Hence, limiting the comparison of Ig levels between intervals to paired data samples decreased the size of the dataset but also reduces the risk selection bias due to singular samples that might not have been followed-up due to confounding reasons. Despite the limitations our analysis provides the first data of different assays and time course of the serological response in a large number of KTR.

## 5. Conclusion

Large individual variability in serological response and loss of serological protection after 4 months in a relevant number of patients supports the utility of regular serological monitoring and might argue for consideration of shorter and individualized vaccination intervals in KTR.

## Data Availability

The data presented in this study are available on request from the corresponding author. The data are not publicly available due to their containing personal information that could compromise the privacy of involved patients.

## Supplementary Materials

**Supplement figure 1.**
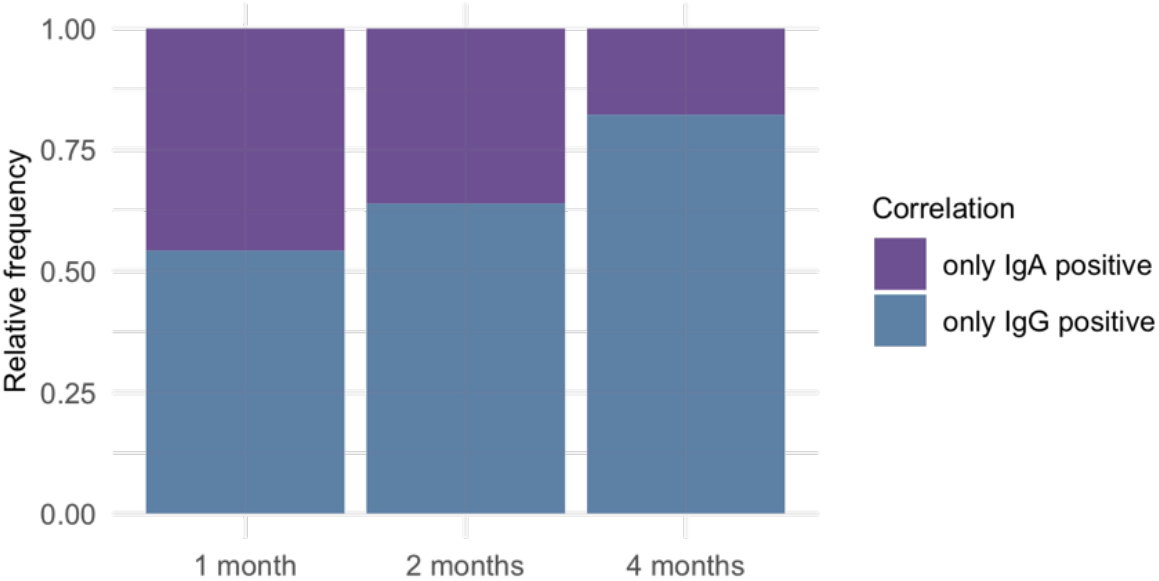
266 cases where IgA and IgG assays determined differing results (only IgG positive vs. only IgA positive measurements) and their relative frequency over time

**Supplement figure 2.**
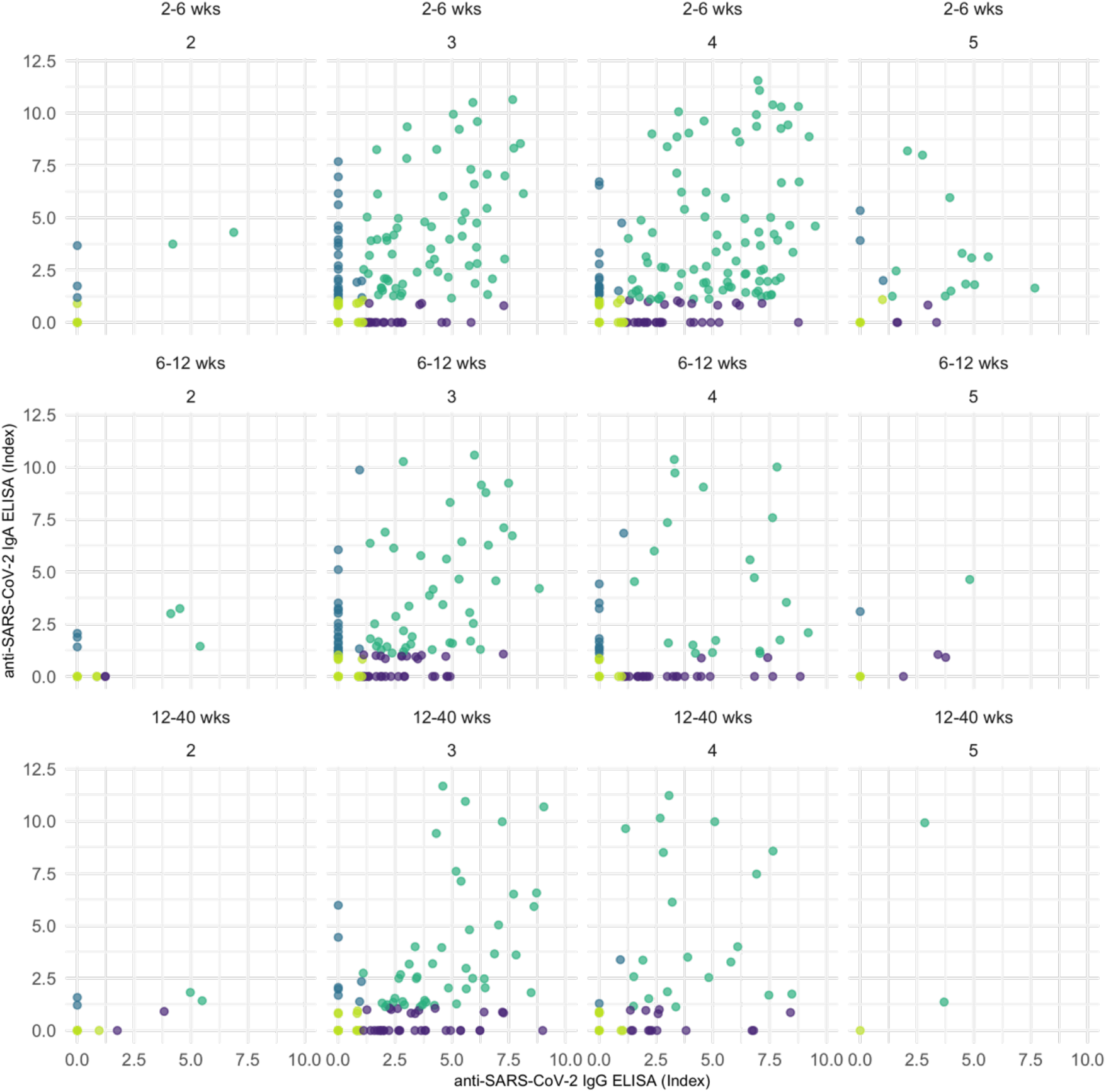
Correlation scatter plots of serological samples performed with anti-SARS-CoV-2 IgG ELISA and IgA ELISA assays. Split by number of vaccine dose (second, third, fourth or fifth vaccination for the individual) and time after vaccination.

## Author Contributions

Conceptualization, Simon Ronicke, Bilgin Osmanodja and Eva Schrezenmeier; Data curation, Simon Ronicke, Bilgin Osmanodja, Bettina Eberspächer, Fritz Grunow and Michael Mikhailov; Formal analysis, Simon Ronicke, Klemens Budde and Bettina Eberspächer; Investigation, Klemens Budde, Annika Jens, Charlotte Hammett, Nadine Koch, Bianca Zukunft, Friederike Bachmann, Mira Choi, Ulrike Weber, Bettina Eberspächer, Jörg Hofmann, Fabian Halleck and Eva Schrezenmeier; Methodology, Eva Schrezenmeier; Project administration, Klemens Budde; Resources, Klemens Budde; Visualization, Simon Ronicke; Writing – original draft, Simon Ronicke and Bilgin Osmanodja; Writing – review & editing, Klemens Budde, Annika Jens, Charlotte Hammett, Nadine Koch, Bianca Zukunft, Friederike Bachmann, Mira Choi, Ulrike Weber, Bettina Eberspächer, Jörg Hofmann, Fritz Grunow, Michael Mikhailov, Fabian Halleck and Eva Schrezenmeier.

All authors have read and agreed to the published version of the manuscript.

## Funding

This research received no external funding.

## Institutional Review Board Statement

The study was conducted in accordance with the Declaration of Helsinki, and approved by the Institutional Ethics Committee of Charité – Universitätsmedizin Berlin (protocol code of the ethics votum: EA1/030/22, date of approval March 10, 2022).

## Informed Consent Statement

Informed consent was obtained from all subjects involved in the study.

## Acknowledgments

E.S. is participant in the BIH-Charité Clinician Scientist Program funded by the Charité – Universitätsmedizin Berlin and the Berlin Institute of Health.

## Conflicts of Interest

The authors declare no conflict of interest.

## Notes

### Competing Interest Statement

The authors have declared no competing interest.

### Author Declarations

institutional ethics committee of Charite - Universitaetsmedizin Berlin, Berlin, Germany, ethics votum EA1/030/22

